# Personalized Machine Learning using Passive Sensing and Ecological Momentary Assessments for Meth Users in Hawaii: A Research Protocol

**DOI:** 10.1101/2023.08.24.23294587

**Authors:** Peter Washington

## Abstract

**Background:** Artificial intelligence (AI)-powered digital therapies which detect meth cravings delivered on consumer devices have the potential to reduce these disparities by providing remote and accessible care solutions to Native Hawaiians, Filipinos, and Pacific Islanders (NHFPI) communities with limited care solutions. However, NHFPI are fully understudied with respect to digital therapeutics and AI health sensing despite using technology at the same rates as other races.

**Objective:** We seek to fulfill two research aims: (1) Understand the feasibility of continuous remote digital monitoring and ecological momentary assessments (EMAs) in NHFPI in Hawaii by curating a novel dataset of longitudinal FitBit biosignals with corresponding craving and substance use labels. (2) Develop personalized AI models which predict meth craving events in real time using wearable sensor data.

**Methods:** We will develop personalized AI/ML (artificial intelligence/machine learning) models for meth use and craving prediction in 40 NHFPI individuals by curating a novel dataset of real-time FitBit biosensor readings and corresponding participant annotations (i.e., raw self-reported substance use data) of their meth use and cravings. In the process of collecting this dataset, we will glean insights about cultural and other human factors which can challenge the proper acquisition of precise annotations. With the resulting dataset, we will employ self-supervised learning (SSL) AI approaches, which are a new family of ML methods that allow a neural network to be trained without labels by being optimized to make predictions about the data itself. The inputs to the proposed AI models are FitBit biosensor readings and the outputs are predictions of meth use or craving. This paradigm is gaining increased attention in AI for healthcare.

**Conclusions:** We expect to develop models which significantly outperform traditional supervised methods by fine-tuning to an individual subject’s data. Such methods will enable AI solutions which work with the limited data available from NHFPI populations and which are inherently unbiased due to their personalized nature. Such models can support future AI-powered digital therapeutics for substance abuse.

## Introduction

### Significance

Methamphetamine (meth) abuse is highly prevalent in Hawaii, especially among Indigenous Pacific People (IPP) [1]. Since the 1980s, Hawaii has been considered as the meth capitol of the U.S. Data from the Pacific Health Analytics Collaborative shows that from 2015-2018, 1.5% of Hawaii residents used meth annually [2]. This is more than twice the national rate of 0.6%. According to the Bureau of Alcohol, Tobacco, Firearms and Explosives (ATF), 71% of all drug cases in Hawaii were related to meth [3]. There are major meth-related disparities between NHFPI and other races in Hawaii, with NHFPI exhibiting elevated rates of illicit substance abuse [4]. According to the CDC, NHPI high school students exhibited lifetime meth use of 7.7%, versus 3.7% in White students, 2.7% in Black students, 3.1% in Asian students, and 5.7% in Hispanic and Latino students5 and these disparities continue into adulthood [5]. Digital interventions powered by AI have the potential to reduce these disparities by aiding clinicians in remotely providing care and monitoring patients between visits, especially populations living in rural areas in Hawaii. Furthermore, such technology could be useful in relapse prevention for those hoping to maintain abstinence. Native Hawaiians have home Internet access at comparable rates to non-Native Hawaiians (58% for Native Hawaiians vs. 55% for non-Native Hawaiians in 2022) [6].

The field of AI-based detection of substance abuse using biometric signals measured by wearables is an active field of research across several research labs globally, as documented in a 2022 review paper by Rumbut et al. [7]. Studies in this field tend to collect prediction labels through remote administration of an ecological momentary assessment (EMA), a methodology where participants are periodically asked to answer questions about their psychiatric or behavioral state while living as usual [8-10]. Unfortunately, prior AI models suffer from clinically unacceptable performance. The primary reason for this lackluster performance is because prior methods attempt to train models using data from many patients, which is the status quo in deep learning due to the requirement of massive datasets to for successful training. In contrast to these prior works, we will develop personalized AI models using a method developed by Dr. Washington’s lab which is capable of learning baseline patterns of human behavior and transferring this knowledge to prediction tasks with very few labeled examples to learn from.

My research lab is particularly qualified to carry out this project, as we are already developing computer science methods to support the personalization of AI models for large and mostly unlabeled data streams, with promising preliminary data and publications in preparation supporting this methodology. As a T1/T2 project (i.e., covering basic discovery and initial human trials), my proposed work is significant both in terms of equitable substance abuse therapeutics but also as a general methodology for clinical and translational research (CTR) in other domains. There are countless situations in healthcare where vast amounts of unlabeled data are collected from a single patient. Annotations for the event of interest (e.g., substance abuse) are frequently sparsely dispersed. The development of predictive supervised models is infeasible in such circumstances, as classical approaches cannot handle the complexity of the data and modern deep learning approaches require vast amounts of data.

### Innovation

To support ML development in situations where vast longitudinal data are collected with minimal human-provided annotations, we propose the development of personalized ML models which are trained solely on an individual’s unlabeled data to learn feature representations which are specific to their baseline temporal dynamics. We are creating a novel method and framework which has never been explored in healthcare, consisting of pre-training neural networks to learn the temporal dynamics of a patient’s biosignals. This method will enable deep networks to be trained using relatively small datasets which would not be possible without the self-supervised approach proposed here. This technique is particularly well-suited for massive datasets with few labels.

The application of personalized AI with a diverse population of persons using substances is unique. NHFPI have been understudied and could benefit from novel treatments to address meth use. While we will apply this technological innovation towards the prediction of meth use, multimodal time-series personalization can be applied to a variety of other biology and health problems where (1) multiple signals are sparsely emitted, (2) the baseline signal patterns are specific to each individual, and (3) it is infeasible to acquire the vast amounts of labels required to train a supervised deep learning model.

This method has the potential to dramatically advance the field of precision healthcare by enabling reliable AI/ML predictions from massive but mostly unlabeled datasets which are trained in a self-supervised manner on data from a single user. This setting of large unlabeled datasets with sparse supervision appears frequently in the field of digital healthcare. Notable examples include passive mobile sensing studies for mental health and wellbeing [11-20], digital therapeutics for children with autism spectrum disorder which record video of the child [21-37], and passive brain sensors for brain-computer interfaces [38-46]. As such, this research protocol can be considered as one of the first tests of a broader emerging paradigm in precision health.

## Methods

### Overview

This protocol was approved by the University of Hawaii Institutional Review Board (IRB) under protocol #2022-01030. Additionally, this study has received further scrutiny and approval via the University of Hawaii’s Data Governance Process under request #230410-3.

The long-term methodological goal of the proposed work is to develop novel AI methodologies for predicting health events (e.g., meth use and cravings) from biosensors in a personalized manner. This technical innovation will be applied towards the detection of drug use events from wearable device sensors. The inputs to the proposed AI models are FitBit biosensor readings and the outputs are predictions of meth use or craving (Figure 1). The developed methods developed can pertain to a variety of biomedical domain areas.

**Figure 1.**
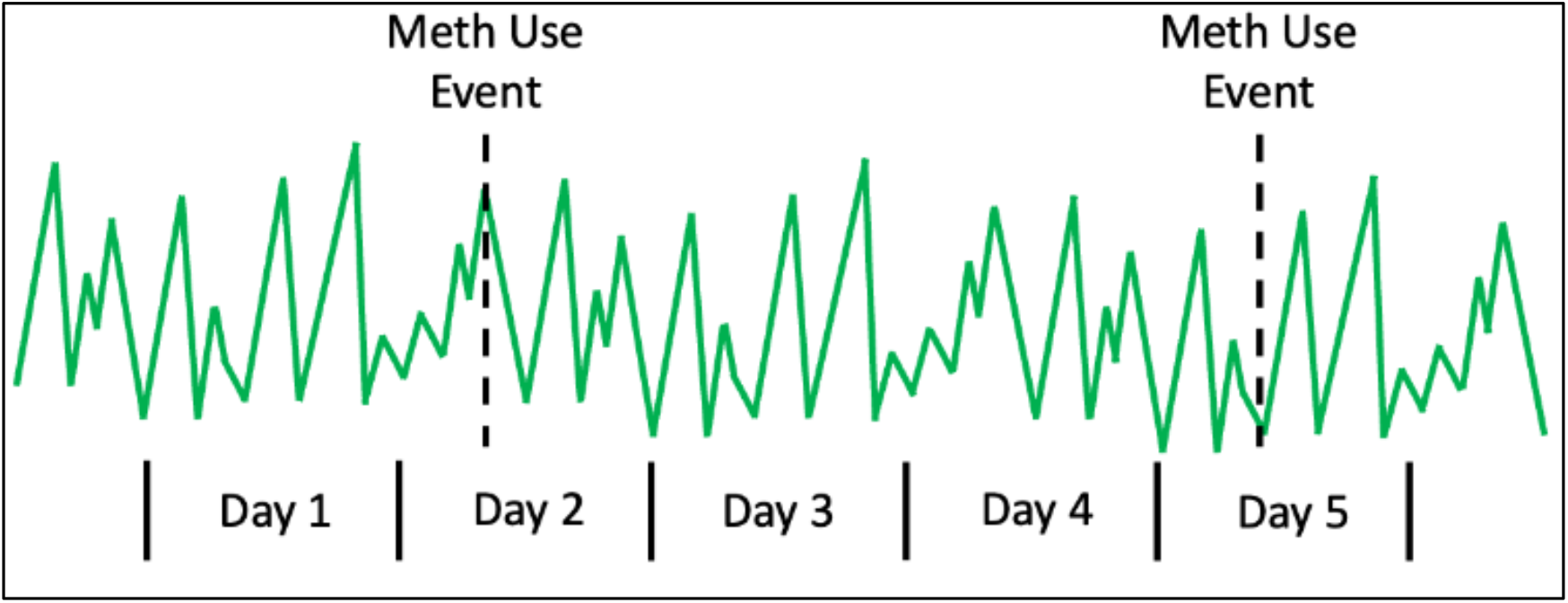
In many biomedical domains, there exist large datasets with sparse annotations of health events. We propose a “personalized self-supervised learning” method which can support training of deep neural networks in such scenarios. We will evaluate our method primarily on the prediction of meth use events using biosignals data from a FitBit.

Diagnostic ML models are typically trained and deployed at a population level. In this traditional scenario, a single model is developed which makes predictions for all individuals within a population. In several health contexts, however, an event of interest occurs repeatedly for an individual. For example, diabetes patients have repeated blood glucose spikes and chronically stressed individuals might have repeated blood pressure spikes. In these cases, ML models can be developed by conducting supervised training on the individual’s data only, resulting in a separate personalized model per individual. While deep learning models have achieved state-of-the-art performance in a variety of health contexts, neural networks require massive datasets which are infeasible to collect for a single individual. However, recent advances in self-supervised learning (SSL), or the subfield of ML focusing on pre-training models without any human-provided labels, are making it possible to realize the personalized ML diagnostics paradigm using deep learning by pre-training the weights of a neural network such that it can learn the baseline temporal dynamics without any labels. The pre-trained model can then transfer learn using relatively few labels that are acquired solely from the individual in question. This methodology can work especially well in scenarios where massive amounts of unlabeled data are collected, such as with continuously worn devices.

**Aim 1: Understand the feasibility of continuous remote digital monitoring and ecological momentary assessments (EMAs) in NHFPI in Hawaii by curating a novel dataset of longitudinal FitBit biosignals with corresponding craving and substance use labels**.

#### Description

We will recruit 40 carefully selected NHFPI participants, who are either in treatment or have received services from one of our community partners, to each participate in a 4-week remote FitBit data collection and concurrent ecological momentary assessment (EMA) study. EMA studies, which involve periodic digital self-reports about psychiatric and behavioral outcomes “in the wild”, have often been used to understand substance abuse [47], including for meth users [48]. We expect >=80% complete data from roughly 25% of the participants (or 10 participants total; see Recruitment section below for justification). Each participant will wear a study-provided FitBit Luxe watch during all waking hours for at least 15 hours each day. Apart from wearing the devices and periodically proving an EMA about their meth use, participants will be asked to follow their normal routine throughout the study.

#### Participant Recruitment and Management

We will recruit participants from a combination of sites, including the Hawai’i Health & Harm Reduction Center (HHHRC) and other sites that the clinical collaborators have connections with (i.e., Hina Mauka). Potential participants will be eligible for the study if they: 1) are 18 years of age or older, 2) self-report consumption of meth on two or more different days per week on average, 3) have no plans to leave Oahu for at least one month, and 4) own a smartphone with either a data plan or regular access to a WiFi connection. Potential participants will be excluded if they: 1) are currently homicidal or suicidal, 2) cannot provide informed consent, 3) are not able to complete interviews in English, 4) expecting incarceration or plan to leave Oahu within the next month, or 5) are unable to provide names and contact information for at least two verifiable locator persons for retention purposes.

We will onboard 40 total participants. A secondary analysis on EMAs for meth abuse monitoring measured the percentage of participants who reached >=80% compliance at different frequencies of meth use, finding that roughly 50% of meth users using 1-3 times per month met this 80% compliance bar, 40% of users using 1-2 days per week met the bar, and 25% of users using 3-4 days per week met the bar [49]. We therefore anticipate a roughly similar compliance rate of around 10 participants (25%) at >=80% compliance, which is sufficient to demonstrate the feasibility of our AI method, as a separate analysis will be conducted for each participant (i.e., 1 model per participant).

#### Data Collection

We will leverage the existing application programming interface (API) provided by FitBit to record the user’s watch sensor readings and upload the data to the cloud. The FitBit API provides access to heart rate (HR), gyroscope, accelerometer, breathing rate, blood oxygen levels (SpO2), and skin temperature sensor readings. These biosensors have previously been used to predict substance abuse and cravings using AI [50-54]. The data will be managed on each participant’s smartphone device through an application, implemented for both iOS and Android, that we are already actively developing. The study team will install the application on the user’s smartphone and configure the FitBit device during study onboarding.

**Table 2.** Study intake/outtake measures and EMA* Questions (adesignates skip logic) collected for each participant.

| Measure/Domain (Time of Administration) | Question |
| --- | --- |
| Background characteristics (Intake) | To better describe the sample, participants will be asked about their sex, gender/sexual orientation, age, race/ethnicity, marital status, education, income, employment, housing, and health insurance status. |
| Substance use and treatment history (Intake) | Questions will be developed to assess current and history of substance use and participation in treatment services. |
| Tolerability (Study conclusion) | How comfortable was the device? |
| Obtrusiveness (Study conclusion) | Did the device change your daily routine? Do you have any concerns with continuously FitBit usage? |
| Meth use (EMA) | Have you used meth since the last time we contacted you?<br><sup>a</sup>Approximately what time did you LAST use meth?<br><sup>a</sup>How did you use meth the last time you used? |
| Craving (EMA) | Please rate your current craving or desire to use meth at this exact moment on a scale of 0-10, with 0 being "no cravings" and 10 being "extremely intense cravings." |

We will run the study with 8 participants at a time, as we have 8 total FitBit Luxe devices, resulting in 5 batches of data collection periods. We will record background characteristics and substance use and treatment history during study intake; we will record questions pertaining to tolerability and obtrusiveness of the app during study outtake (Table 1). The smartphone app will record EMA responses from participants throughout the one-month study period (Table 1). At each EMA, we will ask participants to list approximate times (date, hour, and minute) of their meth intake in the past 24 hours via a user interface on the smartphone app. Participants will be asked to do the same for cravings. Participants will be prompted to provide EMA responses both when drastic signal changes are detected (event-triggered EMA) and every 24 hours (fixed-interval EMA).

We will institute procedures to reduce burden associated with EMA and increase compliance, as suggested by Burke et al [55]. To reduce burden related to the time commitment, we are compensating for every signal-contingent response and providing additional compensation when participants respond to greater than 80% of prompts. Participants will be trained extensively on the EMA protocol during baseline and if they experience any technology-related issues, my research assistants will help troubleshoot remotely. Participants who experience technology-related problems will not have their compensation decreased due to missing prompts.

We will store the curated data from each participant (Figure 2) on a centralized server hosted on Amazon Web Services (AWS). Data uploaded from both wearable systems and the smartphone will first run through a preprocessing server hosted on an Elastic Cloud Compute (EC2) instance with data stored on DynamoDB. Each table will have columns for the participant ID and the timestamp. To ensure privacy and HIPAA compliance, we will encrypt all server-side data and require secret access keys for data access. DynamoDB tables are automatically encrypted on the server side. To add an additional layer of security, we will implement client-side encryption on the mobile application, ensuring encrypted data transmission over an HTTPS connection to move data between the devices and AWS. The data will not be accessible without a secret access key. All data will be anonymized.

**Figure 2.**
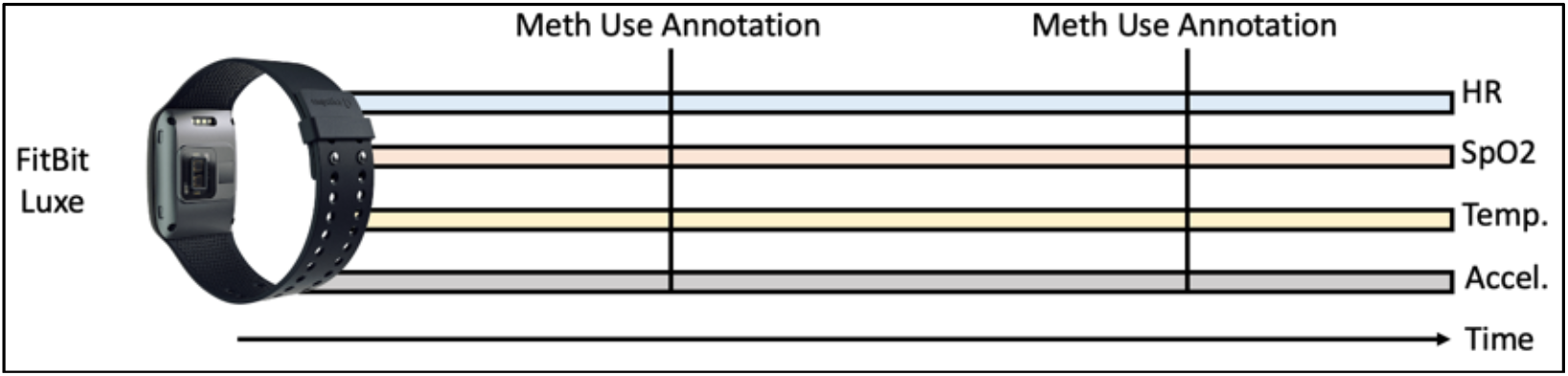
Depiction of the proposed dataset, consisting of continuous FitBit Luxe sensor readings and corresponding meth use and craving annotations from 20 participants collected over a 4-week period.

An anonymized version of participant data will be made available to other computational researchers as a publicly available dataset. This dataset will be stored on Amazon Web Services on a HIPAA-compliant server and will be password protected. Researchers will only gain access to this dataset by signing a Data Use Agreement. Such datasets exist for activity and emotion recognition from wearable data, but prediction of meth use from these measurements will be a challenging task, and other ML practitioners can improve upon our initial AI models with the release of the de-identified dataset. This will be the first publicly available dataset which includes substance use self-reports alongside wearable sensor readings.

#### Data Analysis and Interpretation

We will measure the success rate of remote data collection procedure, as measured by the response rate to EMA notifications. We hypothesize that we will observe higher compliance rates with event-triggered EMAs over fixed interval EMAs. We will also document qualitative challenges with the data collection process, tolerability, and unobtrusiveness (Table 1). We will conduct an interview with participants at the study conclusion when devices are returned. The research team will qualitatively code interview responses to derive recurring themes and design insights.

#### Potential Pitfalls and Mitigation Strategies

This analysis plan is uniquely robust to incomplete data collection because a separate AI model will be trained and evaluated for each participant. There is no requirement for equivalent data streams between participants nor will the analysis be prevented if the full 28-day data collection period is not achieved. The ML strategy can work with only a few logged meth use events. We expect roughly 25% of participants to complete the study at a sufficient level of compliance to support personalized ML analysis. This will provide sufficient data to demonstrate the feasibility of personalized ML analysis.

We have budgeted a 4-month buffer period beyond the 5 months required for complete data collection to account for participants delays and no-shows. Because participant data will be uploaded to AWS daily, we will remotely monitor participants through an automated tracking system already developed by my research lab and will cease the study if compliance is not logged after 4 days. Another possible issue is FitBit theft or loss. To minimize this risk, we will compensate participants $135 for study completion, paid when they return the FitBit. In the case of FitBit device breakage or loss, my lab will purchase additional devices with funds separate from PIKO, up to a limit of 6 additional devices over the course of the study period.

**Aim 2: Develop personalized AI models which predict meth craving events in real time using FitBit sensor data as the model input**.

#### Description

Based on extensive support from prior literature, we hypothesize that AI solutions can detect periods of both meth use and cravings with high sensitivity through personalization of machine learning models. Such models will achieve high performance on a single individual through the fine tuning of each model using only the person of interest’s data for model training. These personalized predictions can trigger the onset of a digital therapy. We will develop two separate AI models per participant: (1) a model which detects meth use in real time, and (2) a model which predicts meth craving in real time. We hypothesize that model personalization using novel self-supervised pre-training strategies will outperform traditional state-of-the-art AI techniques with <5% of the required label data.

#### ML Model Training

The inputs to the models will consist of a separate 1D convolutional backbone pre-trained for each biometric modality. The convolutional features will be fused upstream into a shared joint dense representation space and finally a dense prediction layer with linear activation for regression prediction (Figure 3). We will implement all models using TensorFlow [56].

**Figure 3.**
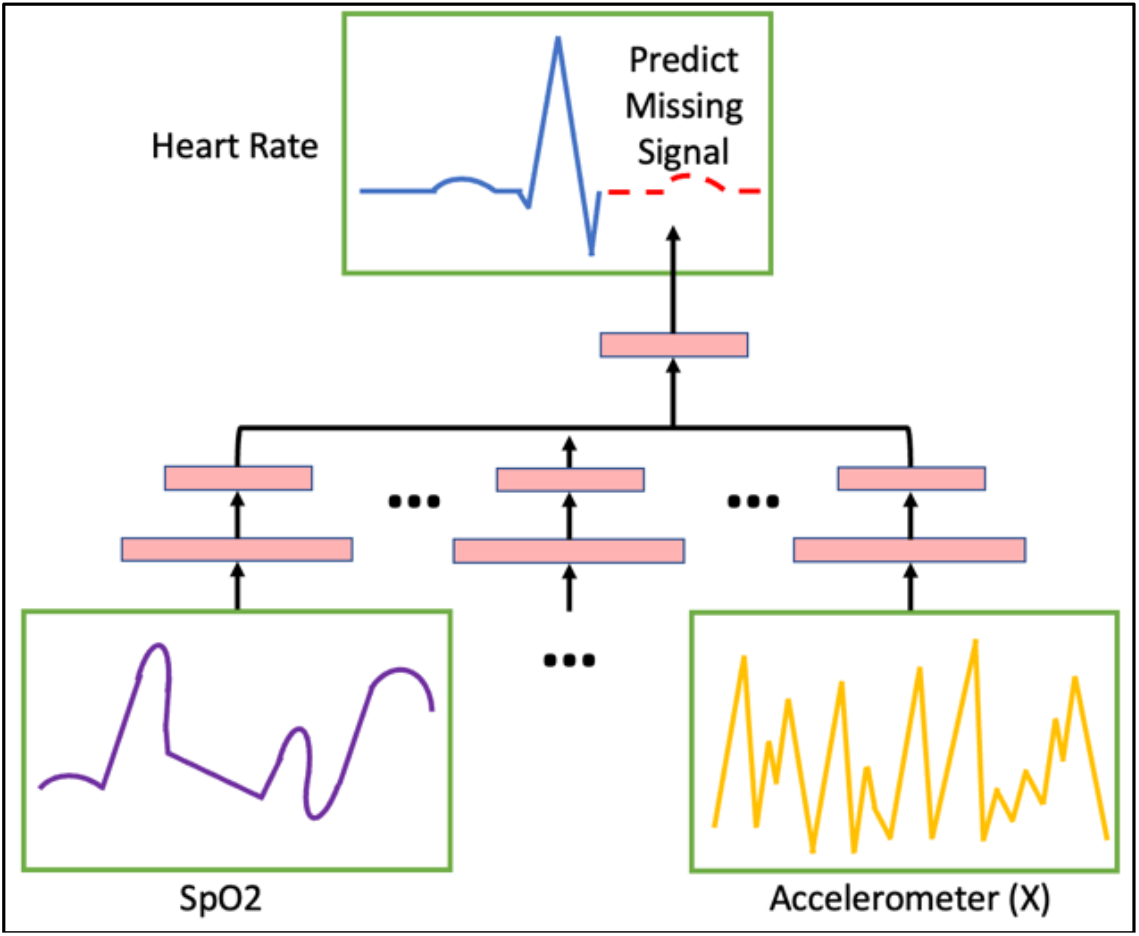
The key methodological computer science innovation of this proposal is the personalization of ML models which make predictions from biosignals time-series data without user-provided labels. Here, we depict a neural network which is trained to predict a heart rate signal given SpO2 and accelerometer signals from a single participant.

The data augmentation techniques that we apply to the signals will be domain-specific, keeping in mind the inherent dynamics of each sensor. For example, for accelerometer data, rotations simulate different sensor placements and cropping is used to diminish the dependency of event locations [57]. Across several modalities, sensor noise can be simulated through scaling, magnitude-warping, and jittering [57]. We will be careful to not apply augmentation strategies which might change the meaning of the underlying signal.

#### Model Personalization

SSL is usually used to pre-train an entire dataset with no explicit labeling by humans to guide the supervision task. We propose to redesign the SSL paradigm towards the task of model personalization. By pre-training a model only on the vast amounts of data curated from a single individual, the weights of the neural network will learn to make predictions using the inherent structure of each participant’s biosignals. This is essential because baseline HR, SpO2, skin temperature, and movement patterns, regardless of stress, will vary drastically across individuals, limiting the performance of general-purpose ML models.

We plan to exploit the multimodal time-series nature of the collected data to perform novel SSL pre-training. We will use one or more signals to predict the value of another signal source (Figure 3). The motivation for this approach is that the biometrics of interest recorded by FitBit are correlated [58-61].

#### Data Analysis and Interpretation

We will train the model on the first 75% of data (by time) and calculate the balanced accuracy, precision, recall, F1-score, and area under the receiver operating characteristic (AUROC) curve on the final 25%. This evaluation pattern mimics real-world use, where a model will be calibrated by a user prior to real-world deployment. It is important to emphasize that we will train and test a separate personalized ML model for each individual (up to 40 separate models).

In a similar nature to our preliminary data, we will evaluate the models by comparing the performance with respect to the number of labeled examples used for supervised fine-tuning. A plot of this comparison will elucidate the number of meth annotations required for model calibration to a single individual. We will create a separate plot for each study participant, as the ML portion of this proposal is testing the personalization of ML models rather than a general-purpose one-size-fits-all ML model which is more typical in ML evaluations.

#### Feasibility

Self-supervised pre-training has been successfully demonstrated in several contexts in computer science and even healthcare [58, 62-63], although never in the personalized context which we will explore except for preliminary results which we have recently published. Multimodal SSL has demonstrated success in prior literature [64-67], although not in the personalized manner in which we will innovate in.

## Data Availability

No data was generated.

## Acknowledgements

The project described was supported by grant number U54GM138062 from the National Institute of General Medical Sciences (NIGMS), a component of the National Institutes of Health (NIH), and its contents are solely the responsibility of the author and do not necessarily represent the official view of NIGMS or NIH.

## References

1. Imperial Sr, Kenneth James. Reducing demand for crystal meth in Hawaii: “Ohana” involvement. Colorado Technical University, 2014.

2. Pasia, Nicole. A look at Hawaii’s underfunded meth epidemic. State of Reform, 2021.

3. Earles, D., and G. DeCosta. “Hawaii Meth Project staff interview (2014).” Honolulu, Hawaii.

4. Monick, Bailey, et al. “Eliminating disparities in youth substance use among Native Hawaiian, Micronesian, and sex and gender minorities: A qualitative needs assessment from interviews with public service leaders.” Psychological Services (2022).

5. 5. Substance Abuse and Mental Health Services Administration. “Behavioral Health Barometer: United States, Volume 5: Indicators as measured through the 2017 National Survey on Drug Use and Health and the National Survey of Substance Abuse Treatment Services.” (2017).

6. 6. Kamehameha Schools’ Strategy & Transformation Group, Liliʻuokalani Trust, Office of Hawaiian Affairs, and Papa Ola Lokahi. ʻImi Pono Hawaiʻi Wellbeing Survey Dashboard. Honolulu: Author, May 2022.

7. Rumbut, Joshua, et al. “An Overview of Wearable Biosensor Systems for Real-Time Substance Use Detection.” IEEE Internet of Things Journal (2022).

8. Ferreri, Florian, et al. “e-Addictology: an overview of new technologies for assessing and intervening in addictive behaviors.” Frontiers in psychiatry (2018): 51.

9. 9. Jacobson, Nicholas C., and Sukanya Bhattacharya. “Digital biomarkers of anxiety disorder symptom changes: Personalized deep learning models using smartphone sensors accurately predict anxiety symptoms from ecological momentary assessments.” Behaviour Research and Therapy 149 (2022): 104013.

10. Tutunji, Rayyan, et al. “Using wearable biosensors and ecological momentary assessments for the detection of prolonged stress in real life.” bioRxiv (2022): 2021–06.

11. 11. Adler, Daniel A., et al. “Machine learning for passive mental health symptom prediction: Generalization across different longitudinal mobile sensing studies.” Plos one17.4 (2022): e0266516.

12. Spathis, Dimitris, et al. “Passive mobile sensing and psychological traits for large scale mood prediction.” Proceedings of the 13th EAI International Conference on Pervasive Computing Technologies for Healthcare. 2019.

13. 13. Lind, Monika N., et al. “The Effortless Assessment of Risk States (EARS) tool: An interpersonal approach to mobile sensing.” JMIR Mental Health5.3 (2018): e10334.

14. Trifan, Alina, Maryse Oliveira, and José Luís Oliveira. “Passive sensing of health outcomes through smartphones: systematic review of current solutions and possible limitations.” JMIR mHealth and uHealth7.8 (2019): e12649.

15. Rabbi, Mashfiqui, et al. “Passive and in-situ assessment of mental and physical well-being using mobile sensors.” Proceedings of the 13th international conference on Ubiquitous computing. 2011.

16. 16. Boonstra, Tjeerd W., et al. “Using mobile phone sensor technology for mental health research: integrated analysis to identify hidden challenges and potential solutions.” Journal of medical Internet research20.7 (2018): e10131.

17. Young, Alexander S., et al. “Passive Mobile Self-tracking of Mental Health by Veterans With Serious Mental Illness: Protocol for a User-Centered Design and Prospective Cohort Study.” JMIR research protocols11.8 (2022): e39010.

18. Wen, Hongyi, et al. “Mpulse mobile sensing model for passive detection of impulsive behavior: Exploratory prediction study.” JMIR Mental Health8.1 (2021): e25019.

19. Wang, Rui, et al. “Tracking depression dynamics in college students using mobile phone and wearable sensing.” Proceedings of the ACM on Interactive, Mobile, Wearable and Ubiquitous Technologies2.1 (2018): 1–26.

20. Wang, Rui, et al. “CrossCheck: toward passive sensing and detection of mental health changes in people with schizophrenia.” Proceedings of the 2016 ACM international joint conference on pervasive and ubiquitous computing. 2016.

21. Banerjee, Agnik, et al. “Training and profiling a pediatric facial expression classifier for children on mobile devices: machine learning study.” JMIR formative research 7 (2023): e39917.

22. Daniels, Jena, et al. “Feasibility testing of a wearable behavioral aid for social learning in children with autism.” Applied clinical informatics9.01 (2018): 129–140.

23. Daniels, Jena, et al. “Exploratory study examining the at-home feasibility of a wearable tool for social-affective learning in children with autism.” NPJ digital medicine1.1 (2018): 32.

24. Kalantarian, Haik, et al. “A mobile game for automatic emotion-labeling of images.” IEEE transactions on games12.2 (2018): 213–218.

25. Kalantarian, Haik, et al. “A gamified mobile system for crowdsourcing video for autism research.” 2018 IEEE international conference on healthcare informatics (ICHI). IEEE, 2018

26. Kalantarian, Haik, et al. “The performance of emotion classifiers for children with parent-reported autism: quantitative feasibility study.” JMIR mental health7.4 (2020): e13174.

27. Kalantarian, Haik, et al. “Labeling images with facial emotion and the potential for pediatric healthcare.” Artificial intelligence in medicine 98 (2019): 77–86.

28. Kalantarian, Haik, et al. “Guess What? Towards Understanding Autism from Structured Video Using Facial Affect.” Journal of healthcare informatics research 3 (2019): 43–66.

29. Kline, Aaron, et al. “Superpower glass.” GetMobile: Mobile Computing and Communications23.2 (2019): 35–38.

30. Penev, Yordan, et al. “A mobile game platform for improving social communication in children with autism: a feasibility study.” Applied clinical informatics12.05 (2021): 1030–1040.

31. Varma, Maya, et al. “Identification of social engagement indicators associated with autism spectrum disorder using a game-based mobile app: comparative study of gaze fixation and visual scanning methods.” Journal of Medical Internet Research24.2 (2022): e31830.

32. Voss, Catalin, et al. “Effect of wearable digital intervention for improving socialization in children with autism spectrum disorder: a randomized clinical trial.” JAMA pediatrics173.5 (2019): 446–454.

33. Voss, Catalin, et al. “Superpower glass: delivering unobtrusive real-time social cues in wearable systems.” Proceedings of the 2016 ACM International Joint Conference on Pervasive and Ubiquitous Computing: Adjunct. 2016.

34. Washington, Peter, et al. “Data-driven diagnostics and the potential of mobile artificial intelligence for digital therapeutic phenotyping in computational psychiatry.” Biological Psychiatry: Cognitive Neuroscience and Neuroimaging5.8 (2020): 759–769.

35. Washington, Peter, et al. “A wearable social interaction aid for children with autism.” Proceedings of the 2016 CHI Conference Extended Abstracts on Human Factors in Computing Systems. 2016.

36. Washington, Peter, et al. “Improved digital therapy for developmental pediatrics using domain-specific artificial intelligence: Machine learning study.” JMIR Pediatrics and Parenting5.2 (2022): e26760.

37. Washington, Peter, et al. “SuperpowerGlass: a wearable aid for the at-home therapy of children with autism.” Proceedings of the ACM on interactive, mobile, wearable and ubiquitous technologies1.3 (2017): 1–22.

38. Ahn, Minkyu, et al. “A review of brain-computer interface games and an opinion survey from researchers, developers and users.” Sensors14.8 (2014): 14601–14633.

39. Al-Hudhud, Ghada, et al. “Analyzing passive BCI signals to control adaptive automation devices.” Sensors19.14 (2019): 3042.

40. Bagheri, Mahsa, and Sarah D. Power. “Simultaneous classification of both mental workload and stress level suitable for an online passive brain– computer interface.” Sensors22.2 (2022): 535.

41. Chen, Wei-Chuan, et al. “A multi-channel passive brain implant for wireless neuropotential monitoring.” IEEE Journal of Electromagnetics, RF and Microwaves in Medicine and Biology2.4 (2018): 262–269.

42. 42. Estepp, Justin R., and James C. Christensen. “Electrode replacement does not affect classification accuracy in dual-session use of a passive brain-computer interface for assessing cognitive workload.” Frontiers in Neuroscience 9 (2015): 54.

43. Park, Seonghun, Chang-Hee Han, and Chang-Hwan Im. “Design of wearable EEG devices specialized for passive brain–computer interface applications.” Sensors20.16 (2020): 4572.

44. Sciaraffa, Nicolina, et al. “Evaluation of a new lightweight EEG technology for translational applications of passive brain-computer interfaces.” Frontiers in Human Neuroscience 16 (2022): 901387.

45. 45. Zander, Thorsten O., et al. “Evaluation of a dry EEG system for application of passive brain-computer interfaces in autonomous driving.” Frontiers in human neuroscience 11 (2017): 78.

46. 46. Zander, Thorsten O., and Sabine Jatzev. “Detecting affective covert user states with passive brain-computer interfaces.” 2009 3rd International Conference on Affective Computing and Intelligent Interaction and Workshops. IEEE, 2009.

47. Shiffman, Saul. “Ecological momentary assessment (EMA) in studies of substance use.” Psychological assessment21.4 (2009): 486.

48. Galloway, Gantt P., et al. “Feasibility of ecological momentary assessment using cellular telephones in methamphetamine dependent subjects.” Substance Abuse: Research and Treatment 1 (2008): SART–S428.

49. Turner, Caitlin M., et al. “Race/ethnicity, education, and age are associated with engagement in ecological momentary assessment text messaging among substance-using MSM in San Francisco.” Journal of substance abuse treatment 75 (2017): 43–48.

50. Carreiro, Stephanie, et al. “Wearable sensor-based detection of stress and craving in patients during treatment for substance use disorder: A mixed methods pilot study.” Drug and alcohol dependence 209 (2020): 107929.

51. Mahmud, Md Shaad, et al. “Automatic detection of opioid intake using wearable biosensor.” 2018 International Conference on Computing, Networking and Communications (ICNC). IEEE, 2018.

52. Natarajan, Annamalai, et al. “Detecting cocaine use with wearable electrocardiogram sensors.” Proceedings of the 2013 ACM international joint conference on Pervasive and ubiquitous computing. 2013.

53. Rumbut, Joshua, et al. “Detecting Kratom Intoxication in Wearable Biosensor Data.” 2019 IEEE/ACM International Conference on Connected Health: Applications, Systems and Engineering Technologies (CHASE). IEEE, 2019.

54. Shi, Ruiqi, et al. “mAAS--A Mobile Ambulatory Assessment System for Alcohol Craving Studies.” 2015 IEEE 39th Annual Computer Software and Applications Conference. Vol. 3. IEEE, 2015.

55. Burke, Lora E., et al. “Ecological momentary assessment in behavioral research: addressing technological and human participant challenges.” Journal of medical Internet research19.3 (2017): e7138.

56. Abadi, Martín, et al. “{TensorFlow}: a system for {Large-Scale} machine learning.” 12th USENIX symposium on operating systems design and implementation (OSDI 16). 2016.

57. Um, Terry T., et al. “Data augmentation of wearable sensor data for parkinson’s disease monitoring using convolutional neural networks.” Proceedings of the 19th ACM international conference on multimodal interaction. 2017.

58. Chowdhury, Alexander, et al. “Applying self-supervised learning to medicine: review of the state of the art and medical implementations.” Informatics. Vol. 8. No. 3. MDPI, 2021.

59. Lan, Xiang, et al. “Intra-inter subject self-supervised learning for multivariate cardiac signals.” Proceedings of the AAAI Conference on Artificial Intelligence. Vol. 36. No. 4. 2022.

60. Li, Yuexiang, et al. “Efficient and effective training of COVID-19 classification networks with self-supervised dual-track learning to rank.” IEEE Journal of Biomedical and Health Informatics24.10 (2020): 2787–2797.

61. Tiu, Ekin, et al. “Expert-level detection of pathologies from unannotated chest X-ray images via self-supervised learning.” Nature Biomedical Engineering (2022): 1–8.

62. Krishnan, Rayan, Pranav Rajpurkar, and Eric J. Topol. “Self-supervised learning in medicine and healthcare.” Nature Biomedical Engineering (2022): 1–7.

63. Shurrab, Saeed, and Rehab Duwairi. “Self-supervised learning methods and applications in medical imaging analysis: A survey.” PeerJ Computer Science 8 (2022): e1045.

64. Akbari, Hassan, et al. “Vatt: Transformers for multimodal self-supervised learning from raw video, audio and text.” Advances in Neural Information Processing Systems 34 (2021): 24206–24221.

65. 65. Lee, Michelle A., et al. “Making sense of vision and touch: Self-supervised learning of multimodal representations for contact-rich tasks.” 2019 International Conference on Robotics and Automation (ICRA). IEEE, 2019.

66. Taleb, Aiham, et al. “Multimodal self-supervised learning for medical image analysis.” International Conference on Information Processing in Medical Imaging. Springer, Cham, 2021.

67. Wang, Luyu, et al. “Multimodal self-supervised learning of general audio representations.” arXiv preprint 2104.12807 (2021).

